# Prevalence of Hyperglycemia in Pregnancy using early, standard and late screening: a study in the Primary Care Health in Lima, Peru

**DOI:** 10.1101/2025.01.26.25321156

**Authors:** S.N. Seclén, Lucy Nelly Damas-Casani, Vicky Delia Cecilia Motta-Montoya, Sandra Lesly Jiménez-Martel, Marlon Yovera-Aldana

**Author notes:** **Corresponding author** (MYA).

## Abstract

**Objective:** To determine the prevalence of hyperglycemia in pregnancy (HIP) based on the timing of screening in pregnant women.

**Materials and Methods:** A cross-sectional study was conducted on pregnant women recruited from February 2019 to August 2022 in four primary care centers in Lima, excluding those with known diabetes. Screening involved an Oral Glucose Tolerance Test with 75 g of glucose at the first contact. It was classified as early if performed before 24 weeks, standard between 24-28 weeks, and late if after 29 weeks. For gestational diabetes mellitus (GDM), we used the IADPSG diagnostic criteria for any gestational age. For diabetes in pregnancy (DIP), glucose levels over 126 mg/dl fasting or over 200 mg/dl at 2 hours post-load were used. We calculated prevalence ratios for GDM and DIP based on the type of screening.

**Results:** We included 4,495 pregnant women with an average age of 29 years. The prevalence of HIP was 14.9%, GDM was 14.2%, and DIP was 0.7%. Early GDM screening showed a 23% higher prevalence compared to GDM screening at 24-28 weeks, adjusted for age (PR 1.23; 95% CI 1.01 – 1.49; p=0.036). No differences were found for DIP across screening types.

**Conclusion:** In pregnant women at four primary centers in Lima, one in seven pregnant women (14.9%) had HIP, GDM was 14.2%, and DIP was 0.7%.. Early screening showed the highest proportion of GDM compared to standard or late screening. Longitudinal studies are needed to validate whether this higher early prevalence impacts maternal-perinatal complications.

## Introducción

Globally, approximately 16.7% of pregnancies present hyperglycemia in pregnancy (HIE), defined as any known pre-existing alteration or first diagnosed during gestation. Of these cases, 80.3% are due to gestational diabetes mellitus (GDM). The prevalence of HIE progressively increases with age and 80% originating in low- to middle-income countries where access to healthcare is limited. [1] HIE is associated with maternal and perinatal complications. [2] Normalizing blood glucose levels in patients with GDM and diabetes in pregnancy (DIP) through lifestyle changes, and in some cases pharmacological treatment, has been shown to reduce maternal-perinatal complications by more than 60%. [3]

In the last two decades, the increase in the prevalence of GDM has ranged from 16% to 127%. This is attributed to the progressive rise in obesity by unhealthy lifestyles, greater governmental efforts to conduct universal screening and a diagnostic threshold criteria diminished[4]. In Peru, the prevalence of overweight and obesity in women of reproductive age is 37% and 28%, respectively[5]. Consequently, nearly 50% of pregnant women have excess weight in the first trimester, making it one of the main risk factors for the development of GDM. [6] Moreover, the new International Association of the Diabetes and Pregnancy Study Groups (IADPSG) criteria, have increased the frequency of GDM by as much as 6 to 11 times [7]. Howewer, the diagnosis and treatment are cost-effective when considering postpartum outcomes for both the mother and the child. [8–10]. The International Federation of Gynecology and Obstetrics (IFGO) proposes different strategies to implement universal screening depending on the resources available in each country. Standard screening in the first trimester includes only fasting glucose; however, it suggests that in high-risk populations, an oral glucose tolerance test (OGTT) with 75 g could be performed at the initial visit, repeating it at 24-28 weeks if the first result is negative. [11] Thus, waiting until that gestational age to initiate treatment could delay a potential improvement in the rate of complications, similar to the preventive action of acetylsalicylic acid to prevent preeclampsia. [12]

Recent studies suggest that abnormal, non-diabetic fasting glucose values in early screening impact complications. [13] Both early hyperglycemia and DIP are associated with worse outcomes than GDM diagnosed by standard methods. [14,15] On the other hand, treating GDM identified by IADPSG criteria using OGTT before 22 weeks in pregnant women with at least one risk factor for GDM, has been shown to reduce neonatal outcomes, although it does not affect those related to hypertensive disorders. [16] However, despite the diversity of criteria for GDM, there is no consensus on first-trimester hyperglycemia, with its nomenclature being variable and complicating comparison between regions. [17]

In Lima-Peru, a study conducted in a reference center using universal screening between 24-28 weeks with IADPSG criteria found a prevalence of 16%. [18] This prevalence, which is close to the global average, could differ if evaluated in primary care health facilities, especially if early OGTT (before 24 weeks) is applied. Therefore, the objective of this study is to determine the prevalence of hyperglycemia during pregnancy in four primary care centers in South Lima at the first contact using OGTT.

## Matherial and Methods

### Study Design and clinical setting

A cross-sectional study was conducted analyzing data from the Gestational Education in Diabetes (GEIDI) program implemented in four maternal and child health centers in South Lima. The GEIDI program is an early diagnosis and educational intervention strategy aimed at pregnant women diagnosed with gestational diabetes through the oral glucose tolerance test (OGTT) during their initial contact. This program was funded by the World Diabetes Foundation (WDF) and executed by a professional and technical team from the Peruvian Juvenile Diabetes Association (ADJ in Spanish) and the María Auxiliadora Hospital. Interventions were carried out at the José Carlos Mariátegui Maternal and Child Center (JCM) in Villa María del Triunfo district, San Genaro Maternal and Child Center (SG) and the Virgen del Carmen Maternal and Child Center (VC) in Chorrillos district and Manuel Barreto Maternal and Child Center (MB) in San Juan de Miraflores district. Pregnant women were recruited from February 2019 to August 2022.

### Population, sample and sampling

Pregnant women attended at the four primary care centers, aged 15 years or older, Peruvian nationality, and having resided in the district for more than six months, were included in the study. Fasting glucose screening is part of the procedures of the Ministry of Health in maternal care, but it was applied for the first time in this population through the oral glucose tolerance test (OGTT) using 75 gr of glucose. Patients with known diabetes mellitus prior to pregnancy, uncertain last menstrual period or not confirmed by ultrasound performed before 24 weeks of gestation, and multiple pregnancies were excluded. The sampling method was non-probabilistic census, accepting all pregnant women who met the eligibility criteria based on the provided data.

### Variables

Hyperglycemia in Pregnancy: For this study, diabetes in pregnancy was defined as having fasting glucose values greater than 126 mg/dl (7.0 mmol/l) or values greater than 200 mg/dl (11.1 mmol/l) at 2 hours, at any gestational age. For gestational diabetes mellitus (GDM), we applied the HAPO criteria, where a diagnosis was positive if fasting glucose was between 92 and 125 mg/dl (5.1 to 6.9 mmol/l), glucose at 1 hour was greater than 180 mg/dl (9.9 mmol/l), and glucose at 2 hours was between 153 and 199 mg/dl (8.5 to 11.0 mmol/l), with at least one positive result being sufficient.

Other Variables: Based on the gestational age during the OGTT, the timing of screening was classified as early (24 weeks or less), standard (24-28 weeks), and late (more than 28 weeks). The year of the OGTT (2019, 2020, 2021, 2022) and age group (less than 18; 18 to 24.9; 25 to 29.9; 30 to 34.9; 35 to 39.9; 40 to 44.9; and 45 or more) were also recorded.

### Procedures

The GEIDI program scheduled an Oral Glucose Tolerance Test with 75 g of anhydrous glucose during the first contact of the pregnant woman with the health center, obtaining three venous samples: baseline, at 1 hour, and at 2 hours. Glucose was measured using the glucose oxidase method, employing a Wiener reagent with a coefficient of variation <2%, following standard guidelines for analysis. An online reporting system was established for laboratory results registration. After diagnosis, clinical management was based on the guidelines from the Ministry of Health. Additionally, educational sessions were provided for nutritional management and tools for glycemic self-monitoring were supplied through the donation of glucose meters. Pregnant women were referred to the Hospital María Auxiliadora for evaluation by endocrinology and gynecology if the maternal-infant health centers lacked the necessary specialty or if the patient required evaluation at a higher-level facility.

For the study, the authors’ team requested the laboratory results data of the screened pregnant women from the GEIDI program coordinators. This information was provided without identifying data such as names or national identity document numbers. Data were evaluated and analyzed from January 15 to March 30, 2024 and included age, date of the OGTT, gestational age, source health center, and the glucose values from the OGTT.

### Analysis plan

The health center of origin, age group, gestational age at the time of the oral glucose tolerance test (OGTT), and the type of screening performed were described using absolute and relative frequencies. Additionally, the proportion of altered glycemia, the total number of glucose measurements, and the type of glycemic alteration were calculated. The demographic characteristics of pregnant women were compared based on the timing of screening: early (less than 24 weeks), standard (24-28 weeks), and late (more than 28 weeks).

Crude and age-adjusted prevalence ratios for the occurrence of GDM or diabetes in pregnancy (DIP) were calculated according to the screening strategy using Poisson regression with robust variances. By the same method, the prevalence ratio for GDM or DIP by age group and year of diagnosis was also calculated. Statistical analysis was performed using Stata version 18.1 (Stata Corp, College Station, Texas, USA), with a significance level set at 0.05 for all hypothesis tests.

### Ethical considerations

This protocol was evaluated by the Ethics Committee of the Universidad Peruana Cayetano Heredia, with registration code 200797 and certificate 646-01-21. Patient confidentiality was maintained through the anonymization of records..

## Results

The study was conducted from February 2019 to August 2022, with a total of 4,710 pregnant women screened. 4,495 met the inclusion and exclusion criteria. From the selected group, 2,590 (57.6%) underwent early screening, 901 (20.0%) standard screening, and 1,004 (22.4%) late screening (**Figure 1**). The restrictions imposed in 2020 due to the COVID-19 pandemic led to a decrease in the screening rate; however, this rate recovered in 2021 and 2022.

**Figure 1.**
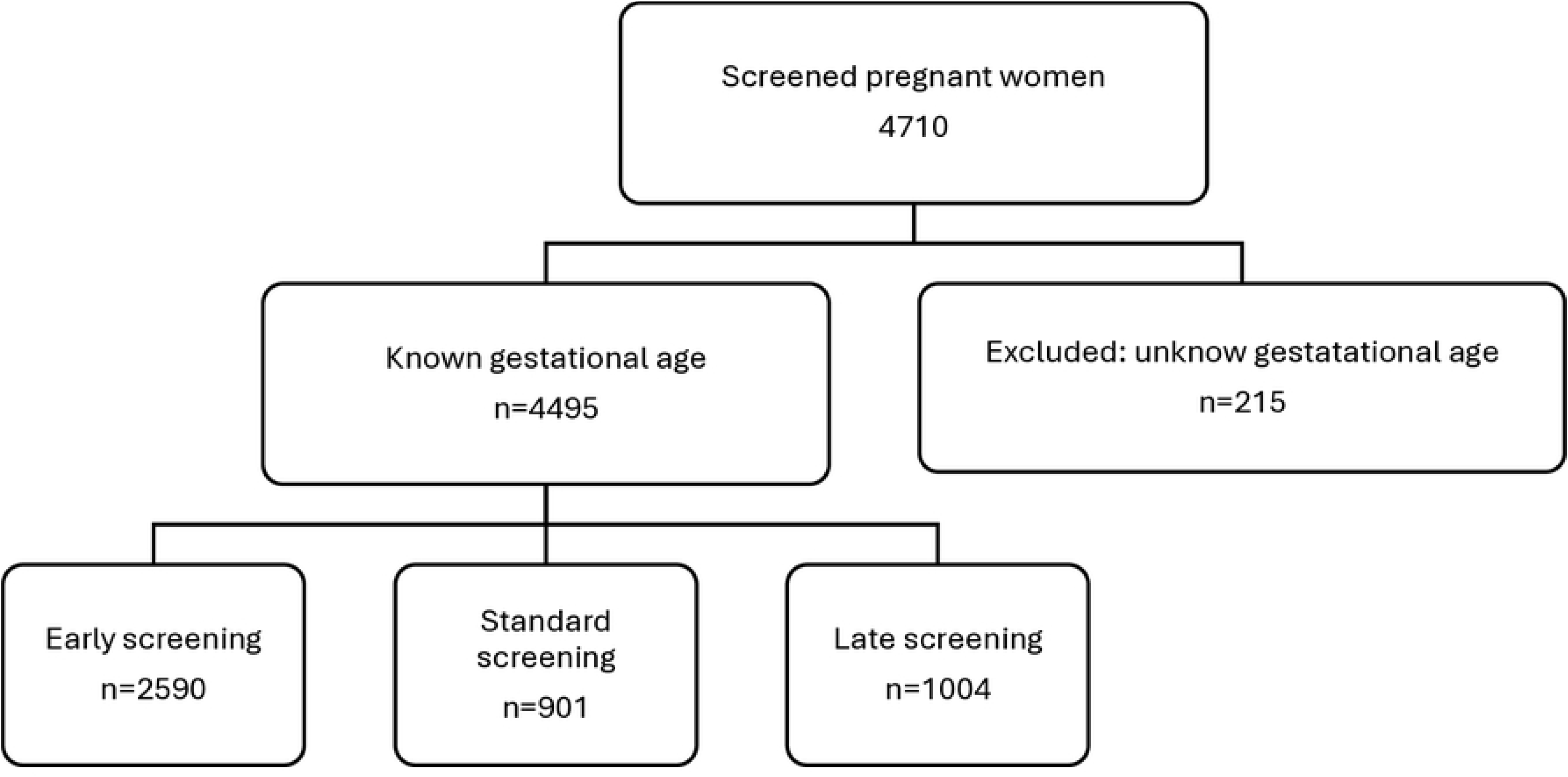
Selection flowchart of the GEIDI Program for the calculation of the prevalence of gestational diabetes mellitus

### General Characteristics

The screened pregnant population was predominantly young, with 78.1% between 18 and 35 years. Regarding OGTT results, abnormal fasting glucose levels were found in 539 pregnant women (12.0%), representing the highest percentage of altered results. Abnormal glucose levels at 1 hour were identified in 193 women (4.2%), and abnormal glucose levels at 2 hours were found in 97 women (2.1%)

Breaking down these findings, it was observed that among the screened pregnant population, only 542 (12.1%) exhibited altered fasting glucose levels, either in isolation or in combination with postprandial glucose levels, and only 127 (2.8%) showed alterations in postprandial glucose levels (**Table 1**).

**Table 1.** Characteristics of pregnant women screened.

| Characterstics | N (%) |
| --- | --- |
| **Total** | 4495 (100.0) |
| **Age (years)** |  |
| Media DE |  |
| < 18 | 121 (2.7) |
| 18 – 24.9 | 1319 (29.3) |
| 25 – 29.9 | 1245 (27.7) |
| 30 – 34.9 | 950 (21.1) |
| 35 – 39.9 | 627 (13.9) |
| 40 – 44.9 | 213 (4.7) |
| 45 and over | 20 (0.4) |
| **Health care facility** |  |
| Jose Carlos Mariátegui VMT | 2400 (53.4) |
| Manuel Barreto SJM | 343 (7.6) |
| San Genaro – Chorrillos | 1233 (27.4) |
| Virgen del Carmen –Chorrillos | 519 (11.6) |
| **Screening year** |  |
| 2019 | 1953 (43.5) |
| 2020 | 657 (14.6) |
| 2021 | 1298 (28.9) |
| 2022 | 587 (13.1) |
| **Screening gestational age** |  |
| 8 or less | 477 (10.6) |
| 9 a 12 | 723 (16.1) |
| 13 a 16 | 559 (12.4) |
| 17 a 23 | 831 (18.5) |
| 24 a 28 | 898 (19.9) |
| 29 a 32 | 492 (10.9) |
| 32 a 36 | 413 (9.2) |
| 37 and over | 102 (2.3) |
| **Screening time** |  |
| Early | 2590 (57.6) |
| Standard | 898 (20.0) |
| Late | 1007 (22.4) |

| **Blood glucose** |  |
| --- | --- |
| Fasting glucose |  |
| Less than 92 mg/dl( 5.1 mmol/l) | 3956 (88.0) |
| 92 to 125.9 mg/dl (5.1 to 6.9 mmol/l) | 520 (11.6) |
| 126 and over (7 mmol/l) | 19 (0.4) |
| Blood glucose at 1 hour |  |
| Less than 180 mg/dl (9.9 mmol/l) | 4302 (95.7) |
| 180 and over (10 mmol/l) | 193 (4.3) |
| Blood glucose at 2 hours |  |
| Less than 153 (8.5 mmol/l) | 4398 (97.8) |
| 153 to 199.9 mg/dl (8.5 to 11.0 mmol/L) | 74 (1.7) |
| 200 and over (11.1 mmol/l) | 23 (0.5) |
| **Type of glycemc alteration** |  |
| No | 3826 (85.1) |
| Basal only | 441 (9.8) |
| Basal and postprandial | 101 (2.3) |
| Postprandial only(s) | 127 (2.8) |

The pregnant women included in the three screening strategies had comparable ages (p=0.213). The early screening strategy exhibited the highest proportion of altered fasting glucose, with 13.5%, compared to 9.9% in the standard screening and 9.8% in the late screening (p=0.010) (**Table 2**).

**Table 2.**
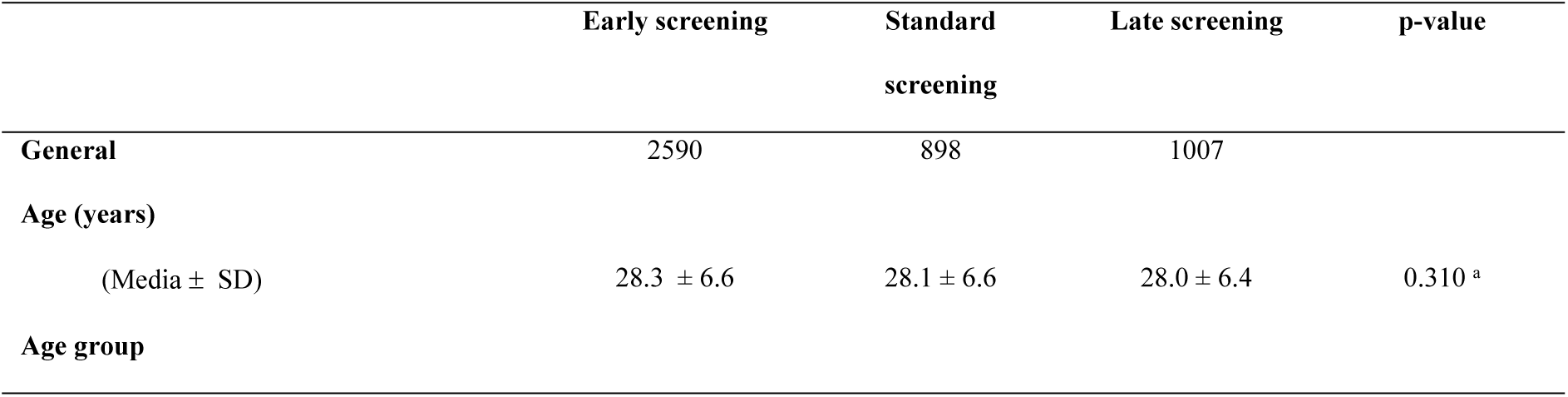
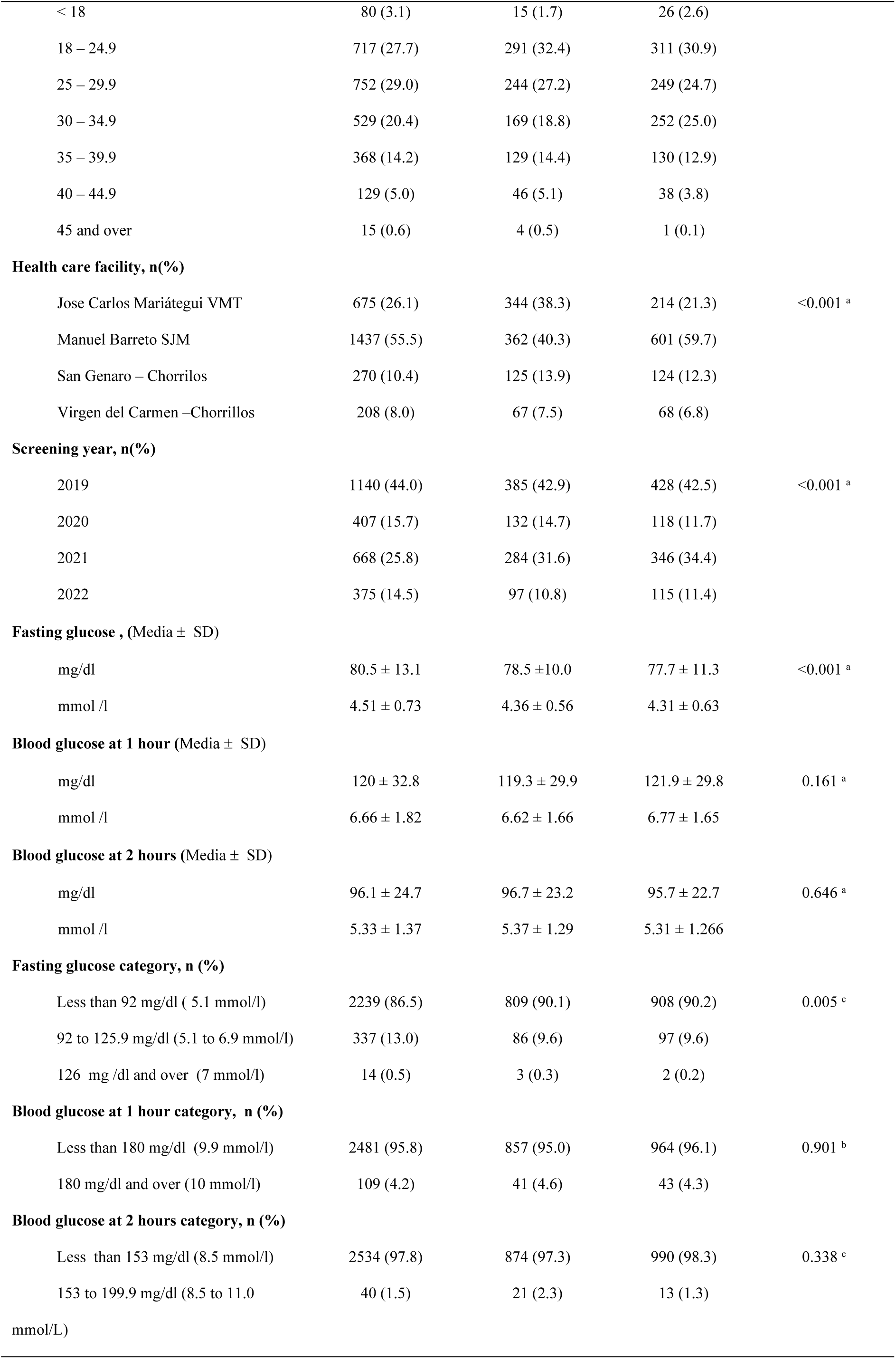
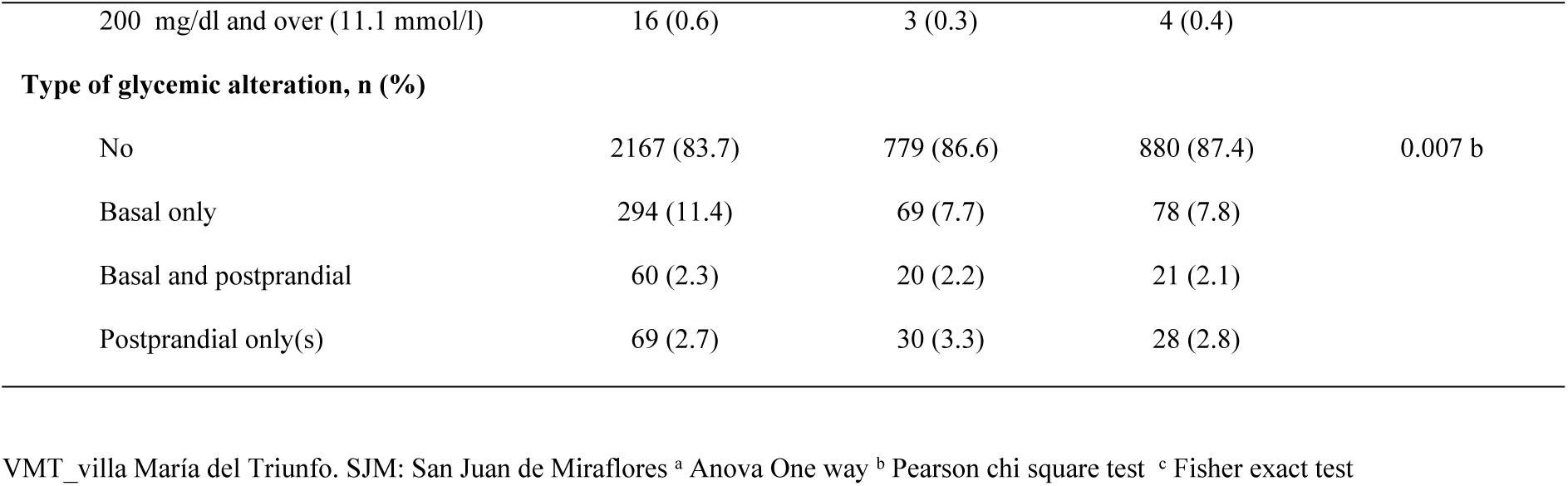
Demographic characteristics according to screening strategy in pregnant women.

|  | Early screening | Standard<br>screening | Late screening | p-value |
| --- | --- | --- | --- | --- |
| **General** | 2590 | 898 | 1007 |  |
| **Age (years)** |  |  |  |  |
| (Media ± SD) | 28.3 ± 6.6 | 28.1 ± 6.6 | 28.0 ± 6.4 | 0.310 <sup>a</sup> |
| **Age group** |  |  |  |  |
| < 18 | 80 (3.1) | 15 (1.7) | 26 (2.6) |  |
| 18 – 24.9 | 717 (27.7) | 291 (32.4) | 311 (30.9) |  |
| 25 – 29.9 | 752 (29.0) | 244 (27.2) | 249 (24.7) |  |
| 30 – 34.9 | 529 (20.4) | 169 (18.8) | 252 (25.0) |  |
| 35 – 39.9 | 368 (14.2) | 129 (14.4) | 130 (12.9) |  |
| 40 – 44.9 | 129 (5.0) | 46 (5.1) | 38 (3.8) |  |
| 45 and over | 15 (0.6) | 4 (0.5) | 1 (0.1) |  |
| **Health care facility, n(%)** |  |  |  |  |
| Jose Carlos Mariátegui VMT | 675 (26.1) | 344 (38.3) | 214 (21.3) | <0.001 <sup>a</sup> |
| Manuel Barreto SJM | 1437 (55.5) | 362 (40.3) | 601 (59.7) |  |
| San Genaro – Chorrillos | 270 (10.4) | 125 (13.9) | 124 (12.3) |  |
| Virgen del Carmen –Chorrillos | 208 (8.0) | 67 (7.5) | 68 (6.8) |  |
| **Screening year, n(%)** |  |  |  |  |
| 2019 | 1140 (44.0) | 385 (42.9) | 428 (42.5) | <0.001 <sup>a</sup> |
| 2020 | 407 (15.7) | 132 (14.7) | 118 (11.7) |  |
| 2021 | 668 (25.8) | 284 (31.6) | 346 (34.4) |  |
| 2022 | 375 (14.5) | 97 (10.8) | 115 (11.4) |  |
| **Fasting glucose , (Media ± SD)** |  |  |  |  |
| mg/dl | 80.5 ± 13.1 | 78.5 ± 10.0 | 77.7 ± 11.3 | <0.001 <sup>a</sup> |
| mmol /l | 4.51 ± 0.73 | 4.36 ± 0.56 | 4.31 ± 0.63 |  |
| **Blood glucose at 1 hour (Media ± SD)** |  |  |  |  |
| mg/dl | 120 ± 32.8 | 119.3 ± 29.9 | 121.9 ± 29.8 | 0.161 <sup>a</sup> |
| mmol /l | 6.66 ± 1.82 | 6.62 ± 1.66 | 6.77 ± 1.65 |  |
| **Blood glucose at 2 hours (Media ± SD)** |  |  |  |  |
| mg/dl | 96.1 ± 24.7 | 96.7 ± 23.2 | 95.7 ± 22.7 | 0.646 <sup>a</sup> |
| mmol /l | 5.33 ± 1.37 | 5.37 ± 1.29 | 5.31 ± 1.266 |  |
| **Fasting glucose category, n (%)** |  |  |  |  |
| Less than 92 mg/dl ( 5.1 mmol/l) | 2239 (86.5) | 809 (90.1) | 908 (90.2) | 0.005 <sup>c</sup> |
| 92 to 125.9 mg/dl (5.1 to 6.9 mmol/l) | 337 (13.0) | 86 (9.6) | 97 (9.6) |  |
| 126 mg /dl and over (7 mmol/l) | 14 (0.5) | 3 (0.3) | 2 (0.2) |  |
| **Blood glucose at 1 hour category, n (%)** |  |  |  |  |
| Less than 180 mg/dl (9.9 mmol/l) | 2481 (95.8) | 857 (95.0) | 964 (96.1) | 0.901 <sup>b</sup> |
| 180 mg/dl and over (10 mmol/l) | 109 (4.2) | 41 (4.6) | 43 (4.3) |  |
| **Blood glucose at 2 hours category, n (%)** |  |  |  |  |
| Less than 153 mg/dl (8.5 mmol/l) | 2534 (97.8) | 874 (97.3) | 990 (98.3) | 0.338 <sup>c</sup> |
| 153 to 199.9 mg/dl (8.5 to 11.0 mmol/L) | 40 (1.5) | 21 (2.3) | 13 (1.3) |  |
| 200 mg/dl and over (11.1 mmol/l) | 16 (0.6) | 3 (0.3) | 4 (0.4) |  |
| **Type of glycemic alteration, n (%)** |  |  |  |  |
| No | 2167 (83.7) | 779 (86.6) | 880 (87.4) | 0.007 b |
| Basal only | 294 (11.4) | 69 (7.7) | 78 (7.8) |  |
| Basal and postprandial | 60 (2.3) | 20 (2.2) | 21 (2.1) |  |
| Postprandial only(s) | 69 (2.7) | 30 (3.3) | 28 (2.8) |  |

VMT\_villa María del Triunfo. SJM: San Juan de Miraflores <sup>a</sup> Anova One way <sup>b</sup> Pearson chi square test <sup>c</sup> Fisher exact test

### Prevalence of Hyperglycemia in Pregnancy

A general prevalence of hyperglycemia in pregnancy was found to be 14.9% (95% CI 13.9-15.9), with gestational diabetes mellitus (GDM) at 14.2% (95% CI 13.2 - 15.3) and diabetes in pregnancy at 0.7% (95% CI 0.5 - 0.9). Of these cases, 95.5% corresponded to GDM (60.2% identified before 24 weeks and 35.3% after 24 weeks), while 4.5% presented with diabetes in pregnancy (**Table 3**).

**Table 3.** Prevalence of hyperglycemia in pregnancy of screened pregnant women in four centers in South Lima.

| Tipo de hiperglicemia en embarazo | Prevalence<sup>a</sup><br>n (%) | CI 95% | Prevalence<sup>b</sup><br>% | CI 95% |
| --- | --- | --- | --- | --- |
| No | 3826 (85.1) |  |  |  |
| Hiperglicemia in pregnancy | 669 (14.9) | (IC95% 13.9 -15.9) | 100% |  |
| Gestational diabetes | 639 (14.2) | (IC95% 13.2 – 15.3) | 95.5% | (IC95% 93.7 – 96.9 ) |
| Early GDM | 403 (8.9) | (IC95% 8.1 – 9.8) | 60.2% | (IC95% 56.4 – 63.9t) |
| Standard GDM | 113 (2.5) | (IC95% 2.1 – 3.0 ) | 16.9% | (IC95% 14.1 -19.9) ) |
| Late GDM | 123 (2.7) | (IC95% 2.2 – 3.2 ) | 18.4% | (IC95% 15.5 – 21.5 ) |
| Diabetes in pregnancy | 30 (0.7) | (IC95% 0.5 - 0.9). | 4.5% | (IC95% 3.0 – 6.3 ) |
| Early DIP | 20 (0.4) | (IC95% 0.2 – 0.6 ) | 3.0% | (IC95% 1.8 – 4.5 ) |
| Standard DIP | 6 (0.1) | (IC95% 0.05 – 0.3 ) | 0.9% | (IC95% 0.3 – 1.9) |
| Late DIP | 4 (0.1) | (IC95% 0.02 – 0.2 ) | 0.6% | (IC95% 0.1 – 1.5) |

GDM: Gestational diabetes DIP: Diabetes n pregnancy
<sup>a</sup> taking as a denominator the total of 4495 pregnant women. <sup>b</sup> Taking as a denominator the totality of 669 pregnant women with HIP

According to the timing of the screening, early showed a higher frequency of gestational diabetes at 26%, compared to standard x(PR 1.26; 95% CI 1.01 – 1.51; p=0.035) (**Table 4**). Tables S2 and S3 provide a description of the patients with GDM and DIP, respectively.

**Table 4.**
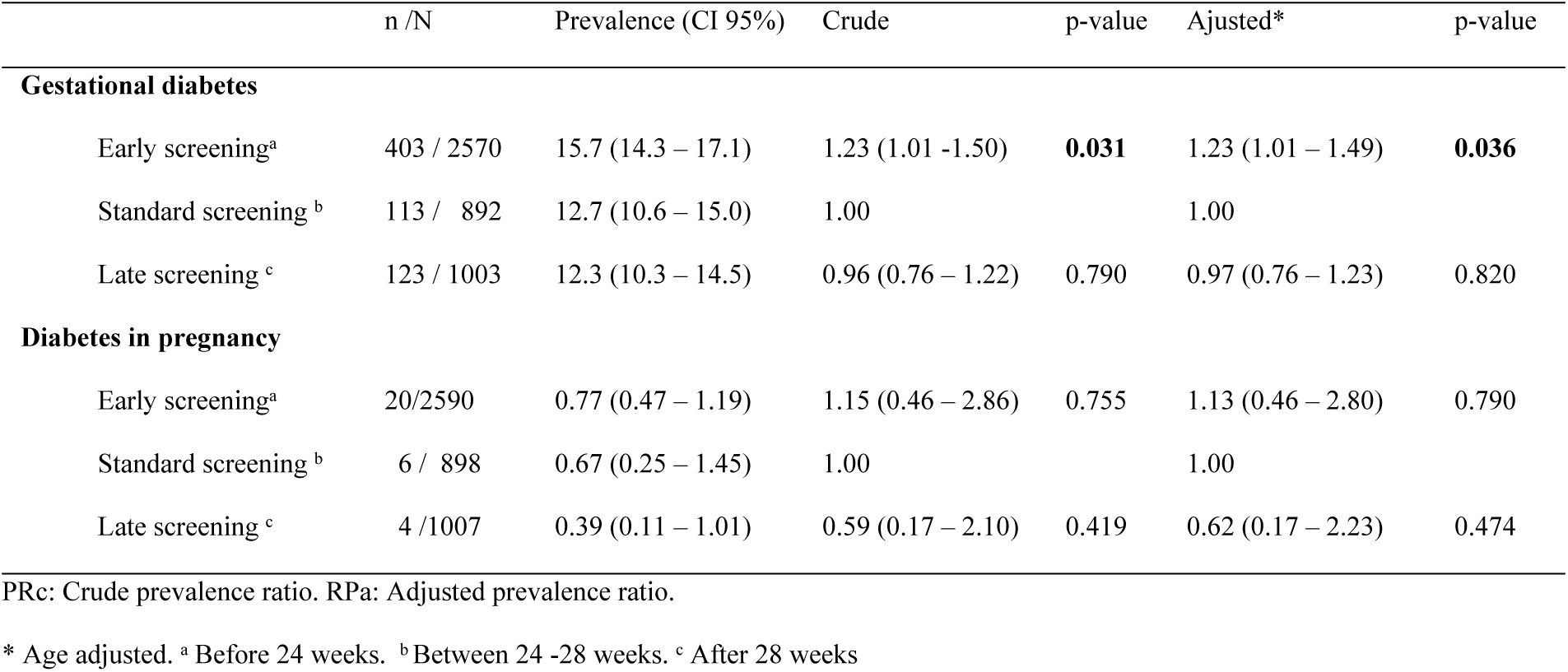
Prevalence ratios of glycemic results according to type of screening.

|  | n /N | Prevalence (CI 95%) | Crude | p-value | Adjusted* | p-value |
| --- | --- | --- | --- | --- | --- | --- |
| **Gestational diabetes** |  |  |  |  |  |  |
| Early screening<sup>a</sup> | 403 / 2570 | 15.7 (14.3 – 17.1) | 1.23 (1.01 -1.50) | **0.031** | 1.23 (1.01 – 1.49) | **0.036** |
| Standard screening <sup>b</sup> | 113 / 892 | 12.7 (10.6 – 15.0) | 1.00 |  | 1.00 |  |
| Late screening <sup>c</sup> | 123 / 1003 | 12.3 (10.3 – 14.5) | 0.96 (0.76 – 1.22) | 0.790 | 0.97 (0.76 – 1.23) | 0.820 |
| **Diabetes in pregnancy** |  |  |  |  |  |  |
| Early screening<sup>a</sup> | 20/2590 | 0.77 (0.47 – 1.19) | 1.15 (0.46 – 2.86) | 0.755 | 1.13 (0.46 – 2.80) | 0.790 |
| Standard screening <sup>b</sup> | 6 / 898 | 0.67 (0.25 – 1.45) | 1.00 |  | 1.00 |  |
| Late screening <sup>c</sup> | 4 /1007 | 0.39 (0.11 – 1.01) | 0.59 (0.17 – 2.10) | 0.419 | 0.62 (0.17 – 2.23) | 0.474 |

PRc: Crude prevalence ratio. RPa: Adjusted prevalence ratio.
\* Age adjusted. <sup>a</sup> Before 24 weeks. <sup>b</sup> Between 24 -28 weeks. <sup>c</sup> After 28 weeks

### Hyperglycemia before 24 weeks

When the OGTT was performed before 24 weeks (early screening), the prevalence of GDM in 2020 showed a decrease of 34% compared to 2019. Subsequently increased a 45% in 2021 and a 92% in 2022. Regarding diabetes in pregnancy (DIP), no significant differences were observed among the years. Additionally, A progressive increase in GDM was evident with advancing age, starting from 30 years. This trend was statistically significant only in the age group of 30 to 40 years (**Table 5**).

**Table 5.**
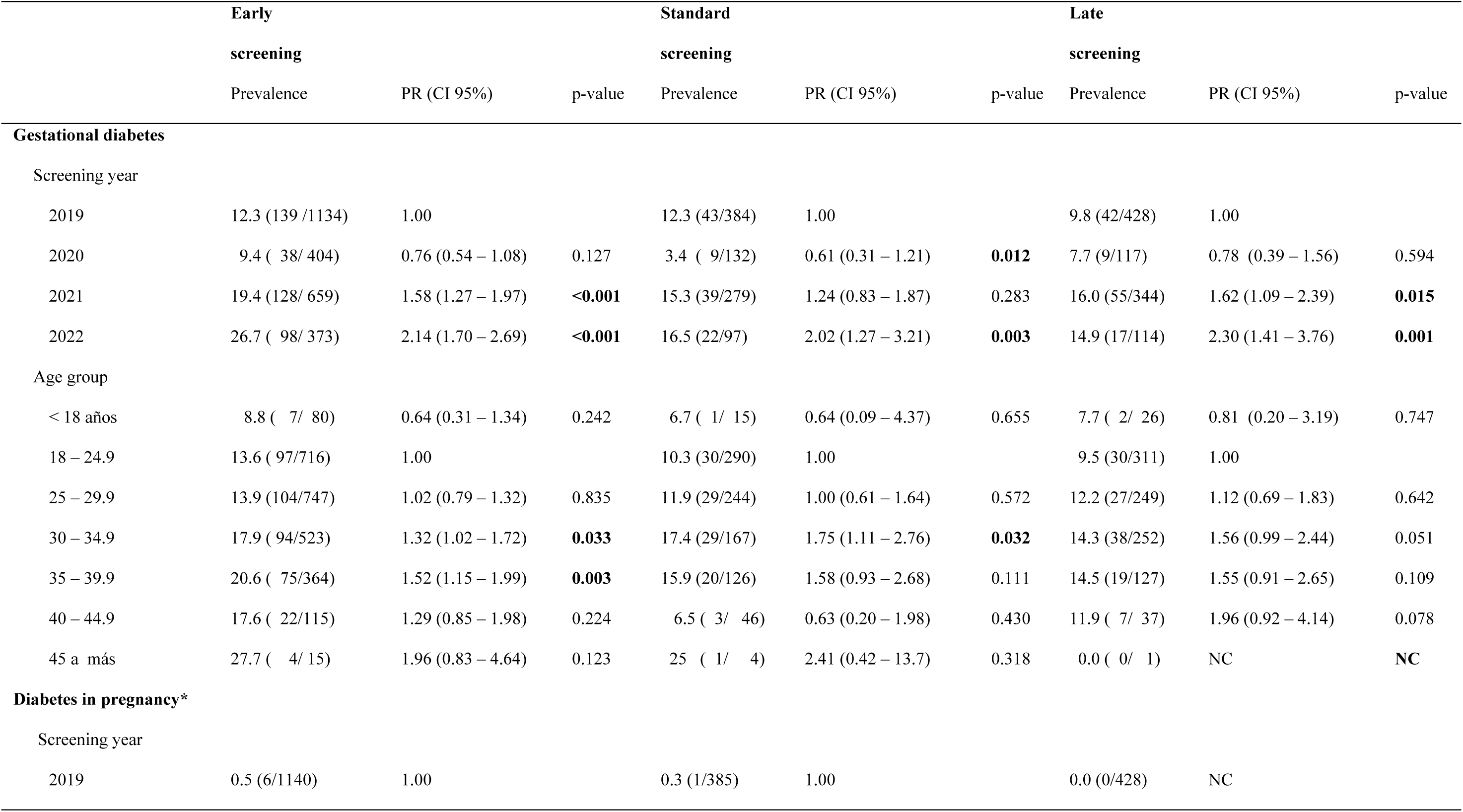
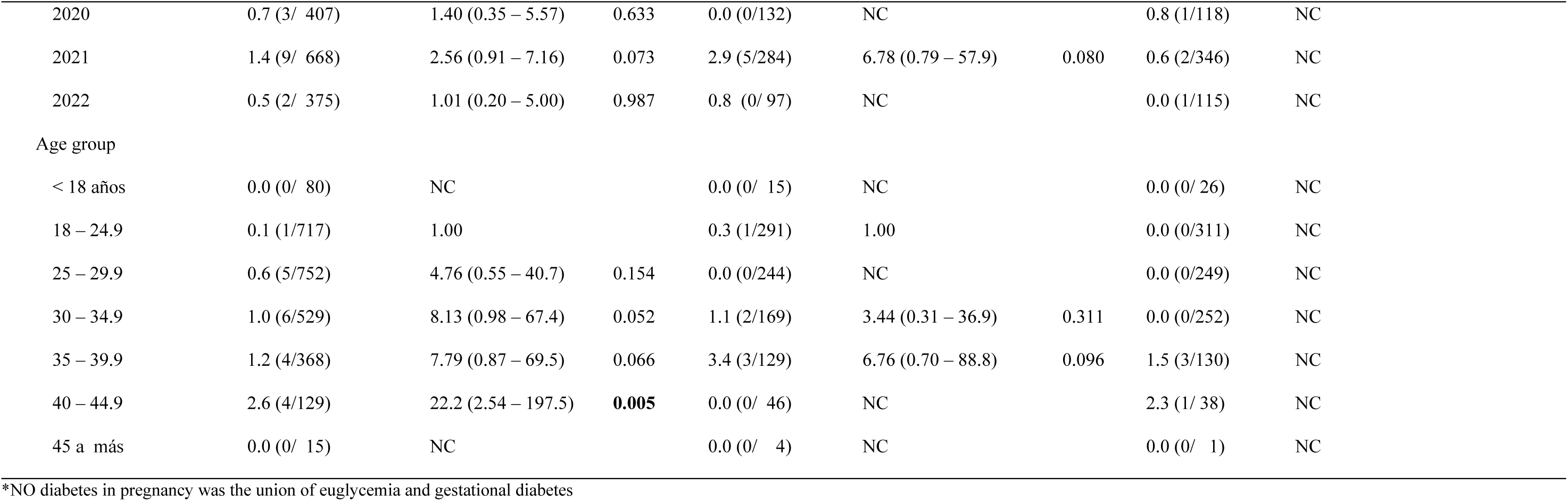
Prevalence ratios of glycemic results according to year, health center and age group of each screening strategy.

|  | Early screening |  |  | Standard screening |  |  | Late screening |  |  |
| --- | --- | --- | --- | --- | --- | --- | --- | --- | --- |
|  | Prevalence | PR (CI 95%) | p-value | Prevalence | PR (CI 95%) | p-value | Prevalence | PR (CI 95%) | p-value |
| **Gestational diabetes** |  |  |  |  |  |  |  |  |  |
| Screening year |  |  |  |  |  |  |  |  |  |
| 2019 | 12.3 (139 /1134) | 1.00 |  | 12.3 (43/384) | 1.00 |  | 9.8 (42/428) | 1.00 |  |
| 2020 | 9.4 ( 38/ 404) | 0.76 (0.54 – 1.08) | 0.127 | 3.4 ( 9/132) | 0.61 (0.31 – 1.21) | **0.012** | 7.7 (9/117) | 0.78 (0.39 – 1.56) | 0.594 |
| 2021 | 19.4 (128/ 659) | 1.58 (1.27 – 1.97) | **<0.001** | 15.3 (39/279) | 1.24 (0.83 – 1.87) | 0.283 | 16.0 (55/344) | 1.62 (1.09 – 2.39) | **0.015** |
| 2022 | 26.7 ( 98/ 373) | 2.14 (1.70 – 2.69) | **<0.001** | 16.5 (22/97) | 2.02 (1.27 – 3.21) | **0.003** | 14.9 (17/114) | 2.30 (1.41 – 3.76) | **0.001** |
| Age group |  |  |  |  |  |  |  |  |  |
| < 18 años | 8.8 ( 7/ 80) | 0.64 (0.31 – 1.34) | 0.242 | 6.7 ( 1/ 15) | 0.64 (0.09 – 4.37) | 0.655 | 7.7 ( 2/ 26) | 0.81 (0.20 – 3.19) | 0.747 |
| 18 – 24.9 | 13.6 ( 97/716) | 1.00 |  | 10.3 (30/290) | 1.00 |  | 9.5 (30/311) | 1.00 |  |
| 25 – 29.9 | 13.9 (104/747) | 1.02 (0.79 – 1.32) | 0.835 | 11.9 (29/244) | 1.00 (0.61 – 1.64) | 0.572 | 12.2 (27/249) | 1.12 (0.69 – 1.83) | 0.642 |
| 30 – 34.9 | 17.9 ( 94/523) | 1.32 (1.02 – 1.72) | **0.033** | 17.4 (29/167) | 1.75 (1.11 – 2.76) | **0.032** | 14.3 (38/252) | 1.56 (0.99 – 2.44) | 0.051 |
| 35 – 39.9 | 20.6 ( 75/364) | 1.52 (1.15 – 1.99) | **0.003** | 15.9 (20/126) | 1.58 (0.93 – 2.68) | 0.111 | 14.5 (19/127) | 1.55 (0.91 – 2.65) | 0.109 |
| 40 – 44.9 | 17.6 ( 22/115) | 1.29 (0.85 – 1.98) | 0.224 | 6.5 ( 3/ 46) | 0.63 (0.20 – 1.98) | 0.430 | 11.9 ( 7/ 37) | 1.96 (0.92 – 4.14) | 0.078 |
| 45 a más | 27.7 ( 4/ 15) | 1.96 (0.83 – 4.64) | 0.123 | 25 ( 1/ 4) | 2.41 (0.42 – 13.7) | 0.318 | 0.0 ( 0/ 1) | NC | **NC** |
| **Diabetes in pregnancy\*** |  |  |  |  |  |  |  |  |  |
| Screening year |  |  |  |  |  |  |  |  |  |
| 2019 | 0.5 (6/1140) | 1.00 |  | 0.3 (1/385) | 1.00 |  | 0.0 (0/428) | NC |  |

|  |  |  |  |  |  |  |  |  |
| --- | --- | --- | --- | --- | --- | --- | --- | --- |
| 2020 | 0.7 (3/ 407) | 1.40 (0.35 – 5.57) | 0.633 | 0.0 (0/132) | NC |  | 0.8 (1/118) | NC |
| 2021 | 1.4 (9/ 668) | 2.56 (0.91 – 7.16) | 0.073 | 2.9 (5/284) | 6.78 (0.79 – 57.9) | 0.080 | 0.6 (2/346) | NC |
| 2022 | 0.5 (2/ 375) | 1.01 (0.20 – 5.00) | 0.987 | 0.8 (0/ 97) | NC |  | 0.0 (1/115) | NC |
| Age group |  |  |  |  |  |  |  |  |
| < 18 años | 0.0 (0/ 80) | NC |  | 0.0 (0/ 15) | NC |  | 0.0 (0/ 26) | NC |
| 18 – 24.9 | 0.1 (1/717) | 1.00 |  | 0.3 (1/291) | 1.00 |  | 0.0 (0/311) | NC |
| 25 – 29.9 | 0.6 (5/752) | 4.76 (0.55 – 40.7) | 0.154 | 0.0 (0/244) | NC |  | 0.0 (0/249) | NC |
| 30 – 34.9 | 1.0 (6/529) | 8.13 (0.98 – 67.4) | 0.052 | 1.1 (2/169) | 3.44 (0.31 – 36.9) | 0.311 | 0.0 (0/252) | NC |
| 35 – 39.9 | 1.2 (4/368) | 7.79 (0.87 – 69.5) | 0.066 | 3.4 (3/129) | 6.76 (0.70 – 88.8) | 0.096 | 1.5 (3/130) | NC |
| 40 – 44.9 | 2.6 (4/129) | 22.2 (2.54 – 197.5) | **0.005** | 0.0 (0/ 46) | NC |  | 2.3 (1/ 38) | NC |
| 45 a más | 0.0 (0/ 15) | NC |  | 0.0 (0/ 4) | NC |  | 0.0 (0/ 1) | NC |

\*NO diabetes in pregnancy was the union of euglycemia and gestational diabetes

### Hyperglycemia after 24 weeks

For the OGTT conducted between 24 and 28 weeks (standard screening), the prevalence of GDM decreased in 2020, followed by increases of 24% and an astonishing 102% in 2021 and 2022, respectively. On the other hand, when the OGTT was performed after 28 weeks (late screening), a higher increase in prevalence was observed, with increases of 62% and 130% for the same years. In standard screening, it was notable that the age group of 30 to 35 years exhibited a 75% increase in prevalence (PR 1.75; 95% CI 1.1-2.76; p=0.032) (**Table 5**).

## Discusión

Hyperglycemia in pregnancy is a condition that significantly impacts maternal and perinatal outcomes, and its screening in primary care is essential. In this study, we found that approximately 1 in 7 pregnant women (14.9%) presents this condition, excluding those with known diabetes prior to pregnancy. Of these cases, 95.5% corresponded to gestational diabetes mellitus (GDM), with 60.2% identified before 24 weeks and 35.3% after this period; additionally, 4.5% presented diabetes in pregnancy.

The global prevalence of subtypes of hyperglycemia in pregnancy varies according to the specific type and geographic region. According to the medical literature, hyperglycemia in pregnancy affects approximately 16.9% of live births worldwide. Within this group, it is estimated that 16.0% of cases are attributed to diabetes in pregnancy, which includes both known diabetes and previously undiagnosed cases.[19] Regarding gestational diabetes, it is estimated to affect approximately 14% of pregnancies worldwide, although this figure can vary depending on risk factors and diagnostic methods used.[20] For instance, in Australia, the prevalence of hyperglycemia in pregnancy was reported at 13.1%, with 12.7% attributed to gestational diabetes and 0.4% to diabetes in pregnancy.[21] The study by Cosson et al. provides data on the prevalence of different subtypes of hyperglycemia in pregnancy, reporting 10.8% for early diagnosed gestational diabetes and 11.7% for later diagnosed gestational diabetes, while diabetes in pregnancy and early diagnosed diabetes have a prevalence of 0.6% each.[15] Furthermore, a systematic review aimed at comparing the prevalence of GDM according to the new IADPSG criteria versus the previous WHO criteria found that early screening increased prevalence by 60% and standard screening by 78%.[22] It is important to highlight that more than 90% of cases of hyperglycemia during pregnancy occur in low- and middle-income countries, underscoring the relevance of this condition from a public health and maternal-infant health perspective, particularly in developing countries.[19]

### Explanation of Results

Several institutions agree that pregnant women with risk factors for diabetes should undergo early screening due to the high associated complication rates.[23] The IADPSG guidelines, adopted by the WHO in 2013, recommended using the same cut-off points between 24-28 weeks at any time during pregnancy.[24] However, it is crucial to differentiate between two conditions that may be detected during early screening: diabetes in pregnancy (pre-gestational) and other lesser hyperglycemic states that might later qualify as GDM but could merely be transient hyperglycemia during the first trimester.

In 2015, an IADPSG panel suggested that fasting glucose values between 92 and 125 mg/dl should not be used for early screening, although they did not offer clear alternatives.[25] This recommendation was based on an Italian study showing that only 45% of patients with fasting glucose greater than 92 mg/dl during the first trimester were positive in the OGTT between 24-28 weeks, with an area under the curve (AUROC) of 0.614. [26] Similarly, a study in China recommended using a cut-off of 110 mg/dl instead of 92 mg/dl for the first trimester, showing a slight but consistent decline in fasting glucose from the first to the second trimester across the entire cohort.[27] In Simmons’ clinical trial, only 67% of pregnant women with early GDM tested positive again in the OGTT at 24 weeks. [16] However, a fasting glucose level greater than 92 mg/dl may be considered a risk factor for the subsequent development of GDM, similar to pregestational body mass index.

The main goal of screening is early detection to reduce the burden of subsequent disease, and not merely to diagnose prematurely without effects on relevant clinical outcomes. It has been documented that early GDM is associated with a higher risk of perinatal mortality, neonatal hypoglycemia, and increased insulin use compared to standard screening.[28] Nevertheless, it remains unclear whether the treatment of early gestational diabetes and its intensive management can be applied to all pregnant women or only those with certain risk factors.[29,30]

This group of “early GDM” may represent a different range of phenotypes compared to those diagnosed after 24 weeks. A study comparing early GDM (before week 21) with late GDM (from week 24 onwards) found lower insulin sensitivity in the early GDM group, with similar beta cell dysfunction. However, the early screening group presented a higher pregestational body mass index (BMI).[31] It is important to discuss the impact of COVID-19, which appeared in the second year of recruitment, leading to a decrease in screening tests. However, when conditions improved, the program continued to follow the same algorithm, resulting in a notable increase in the prevalence of hyperglycemia, reaching up to 60% compared to 2019. Other studies have reported an increase ranging from 14% to 34%. [32,33] A possible explanation for this increase could be related to the rise in pregestational weight and gestational weight gain, which is linked to lifestyle changes such as reduced physical activity and increased stress, as well as changes in healthcare access and service utilization.[34–36]

### Research Recommendations

Longitudinal studies or cost-effectiveness analyses are needed to evaluate the impact on perinatal outcomes for patients diagnosed with early gestational diabetes (GDM) or through standard screening to determine whether this trend is also observed in Latino populations. Furthermore, greater clarity is needed on whether early screening could be implemented universally or based on risk factors (such as obesity or previous history of GDM). It is essential to define the exact timing during the first trimester, considering the physiological changes in pregnant women (before or after 16 weeks) or to use less strict diagnostic glycemic thresholds (recommended 110 mg/dl fasting). Regarding the diagnostic method, the oral glucose tolerance test (OGTT) and its variants are universally accepted. However, they are not easily applied in logistical terms for screening. Alternative diagnostic methods are being explored that offer better feasibility, such as hemoglobin A1c with a cut-off of 5.9%.[37] Additionally, multiple biological markers, such as C-reactive protein, adiponectin, and tumor necrosis factor, are being evaluated either in isolation or in combination with clinical variables as alternatives to the OGTT.[38]

### Limitations and Strengths

Several limitations were identified in the present study. The cut-off points used for early gestational diabetes (GDM) lack long-term studies to support their impact and are based on IADPSG criteria. This may lead to an overestimation of GDM prevalence. Nonetheless, the World Health Organization (WHO) continues to recommend their use. Furthermore, the database did not include other variables that could explain differences in diagnostic prevalence, such as pregestational body mass index or previous history of gestational diabetes. However, we have sufficient statistical power to estimate differences between age groups. The COVID-19 pandemic also affected data collection and the performance of tests. Nevertheless, the tests that were conducted adhered to strict biosafety protocols. It is possible that GDM between 24 and 28 weeks is underrepresented, as the OGTT was not repeated for patients who tested negative in the early screening. Among the strengths of this study, we highlight the sample size, which allows for prevalence estimates with an accuracy of 1.3%. Additionally, the OGTT was used, recognized as the gold standard for diagnosing gestational diabetes.

## Conclusion

In pregnant women from four primary care centers in South Lima, it was found that one in seven (14.9%) presented hyperglycemia in pregnancy. GDM was 14.2%, and DIP was 0.7%. Early screening showed the highest proportion of GDM compared to standard or late screening. No differences in the prevalence of diabetes in pregnancy were found between the groups. Longitudinal studies are needed to determine whether this higher prevalence associated with early screening has repercussions on maternal-perinatal complications and whether it requires any form of treatment.

## Data Availability

All relevant data are within the manuscript and its Supporting Information files.

## Supplementary tables

**Table S1.** Results of basal glycemia, 60 minutes and 120 minutes of OGTT according to clinical characteristics in pregnant women

**Table S2.** Characteristics of patients with gestational diabetes

**Tabla S3.** Carácterísticas de las pacientescon Diabetes pregestacional.

## Notes

### Competing Interest Statement

The authors have declared no competing interest.

### Funding Statement

The author(s) received no specific funding for this work.

